# DeepFLAIR*: Improving Multiple Sclerosis Diagnostic Imaging Workflow Using Deep Learning

**DOI:** 10.1101/2025.11.25.25340993

**Authors:** Inga Baburyan, Bryan Quah, Sreekanth Madhusoodhanan Nair, Omar Al-Louzi, Marcel Maya, Marwa Kaisey, Nancy L. Sicotte, Jason H. Moore, Daniel Ontaneda, Pascal Sati

**Author notes:** The authors declare no competing interests. **Correspondence:** Inga Baburyan, PhD Candidate.

## Abstract

**Background:** Magnetic resonance imaging (MRI) plays a central role in diagnosing multiple sclerosis (MS), yet conventional T2-FLAIR imaging provides limited specificity for distinguishing MS lesions from other white matter abnormalities. The Central Vein Sign (CVS) is a sensitive and specific imaging biomarker which was recently included in the 2024 McDonald criteria for MS diagnosis. FLAIR*, which combines T2-FLAIR and T2* 3D EPI acquisitions, provides optimal detection of the CVS; however, this post-processing workflow requires two separate scans which increases scan time, susceptibility to motion artifacts, and registration error, thus limiting clinical deployment. This study aims to address this issue using a novel deep learning methodology called DeepFLAIR*.

**Methods:** Retrospective analysis was performed on multicenter 3-Tesla brain MRI data as part of the Central Vein Sign in Multiple Sclerosis (CAVS-MS) study. The dataset included 315 participants scanned on Siemens and Philips 3T systems using standardized protocols incorporating 3D T2-FLAIR and 3D T2*-weighted EPI acquisitions (0.65-mm isotropic resolution; scan times ≈ 6-7 minutes per sequence). A 3D U-Net-based conditional generative model, DeepFLAIR*, was developed to synthesize FLAIR* contrast directly from single-sequence T2* 3D EPI images. The model was trained and validated using 89 subjects and tested on an independent cohort of 226 subjects. Quantitative evaluation included structural similarity index (SSIM), peak signal-to-noise ratio (PSNR), mean squared error (MSE), and contrast-to-noise ratio (CNR) across lesion-vein, lesion-white matter, vein-white matter, and white matter-cerebrospinal fluid regions. Statistical comparisons between real-world and synthetic FLAIR* images were performed using paired Wilcoxon signed-rank tests with false discovery rate correction (α = 0.05).

**Results:** Quantitative metrics confirmed that DeepFLAIR* achieved significantly improved contrast-to-noise ratios and comparable global similarity measures relative to real-world FLAIR* (P < 0.001). Synthetic FLAIR* images demonstrated high structural fidelity to real-world FLAIR* (SSIM = 0.78 ± 0.03, PSNR = 23.6 ± 1.35 dB, MSE = 0.0045 ± 0.0015). CNR analyses revealed enhanced lesion-vein and vein-white matter contrast, confirming preservation of perivenular morphology relevant to CVS detection. Lesion morphology and vein-lesion spatial relationships were consistently preserved across subjects.

**Conclusions:** This study demonstrates feasibility of our novel DeepFLAIR* methodology for generating diagnostically relevant FLAIR* contrast from a single T2* 3D EPI input, thereby eliminating the need for dual acquisitions and offline post-processing. This approach could streamline MRI workflows, expand clinical access to CVS-based MS evaluation, and facilitate automated biomarker detection in future diagnostic pipelines.

## INTRODUCTION

Multiple sclerosis (MS) is a chronic inflammatory disease of the central nervous system and a leading cause of neurological disability in young adults^1–3^. Magnetic resonance imaging (MRI) has become instrumental in establishing a diagnosis in individuals at first clinical presentation. Using conventional T2-FLAIR and T1 post-contrast brain imaging, dissemination in space (DIS) can be demonstrated by the presence of hyperintense T2 lesions in two or more key areas of the CNS, and dissemination in time (DIT) can be demonstrated by the presence of at least one T1-enhancing lesion or new hyperintense T2 lesion on a follow-up MRI^4,5^. While conventional MRI is highly sensitive in detecting white matter lesions, it lacks specificity for distinguishing MS lesions from non-specific white matter changes related to migraine and small vessel disease^4,5^. This lack of specificity is particularly problematic in real-world clinical practice where MS misdiagnosis has been recently estimated to affect approximately one in five MS patients, often subjecting the patient to unwarranted exposure to disease-modifying therapies (DMTs), unnecessary healthcare utilization, and psychological distress^6^.

Recent imaging advances have emphasized the diagnostic value of perivenular lesion morphology. The Central Vein Sign (CVS), defined as the presence of a small vein traversing the core of a white matter lesion, has demonstrated high diagnostic sensitivity (92%) and specificity (82%)^7,8^. Accordingly, the presence of CVS can accurately differentiate MS from other diseases and was incorporated into the 2024 revisions of the McDonald criteria for MS diagnosis^9^. These updates highlight the increasing importance of imaging methods that reliably visualize perivenular lesion architecture.

FLAIR* imaging, obtained by combining T2-FLAIR (7-minute scan time) and T2*-weighted 3D-EPI (6-minute scan time) scans, is an optimized contrast for the detection of CVS^10,11^ which accurately differentiate MS lesions from non-MS lesions^8,12–18^. However, it currently requires two separate acquisitions and offline post-processing, which prevents real-time clinical access, increases scan time, and is highly susceptible to motion artifacts and accurate image registration parameters. These workflow limitations can restrict routine implementation in the clinical MRI settings.

To overcome these barriers, we developed DeepFLAIR*, a 3D conditional generative network model that synthesizes FLAIR* images directly from a single T2* 3D EPI input, eliminating the need for dual-sequence acquisition and post-hoc image fusion. DeepFLAIR* is designed to replicate the combined lesion- and cerebrospinal fluid (CSF)-suppressed contrast of true FLAIR*, enabling direct visualization of perivenular lesions for MS diagnosis. In this study, we evaluated DeepFLAIR* in terms of whole-brain image similarity, lesion conspicuity, and preservation of CVS visibility relevant to MS diagnostic interpretation.

## METHODS

### Data Acquisition

MR imaging dataset of two cohorts from the Central Vein Sign: A Diagnostic Biomarker in Multiple Sclerosis (CAVS-MS) multicenter studies were used^14,19^. Institutional Review Board approval was obtained by Cleveland Clinic. A pilot cohort (N_train=89) was used for model training through hyper-parameter search and model selection, while a larger cohort (N_test=226) was used for model testing and evaluation (total N = 315; mean age = 41.3 years; 239 females, 76 males). Subjects were scanned using standardized imaging protocols including T2-FLAIR and T2* 3D-EPI acquisitions on Siemens and Philips 3T MRI systems. Real-world FLAIR* volumes were generated via post-processing of paired T2-FLAIR and T2* 3D-EPI images through linear image registration, interpolation, and multiplication of the two separate scans^10^.

### Data Processing and Deep Learning Architecture

The proposed network, DeepFLAIR*, consisted of a 3D U-Net–based image-to-image translation model to synthesize FLAIR* volumes from single-sequence T2* 3D EPI inputs^20^ (**Figure 1A**). The training set of T2* input volumes was intensity-normalized to the range [0, 1] on a per-volume basis and padded to a uniform size of 320×384×320 voxels. For model training, data augmentation included image flipping (horizontal and/or vertical) and image blurring using a Gaussian filter (none, mild, or strong) was performed. Overlapping 3D patches of size 64×64×64 voxels were extracted using a stride of 32, yielding a consistent patch count per subject. For model testing, images were preprocessed in a similar manner without data augmentation (**Figure 1B)**.

**Figure 1:**
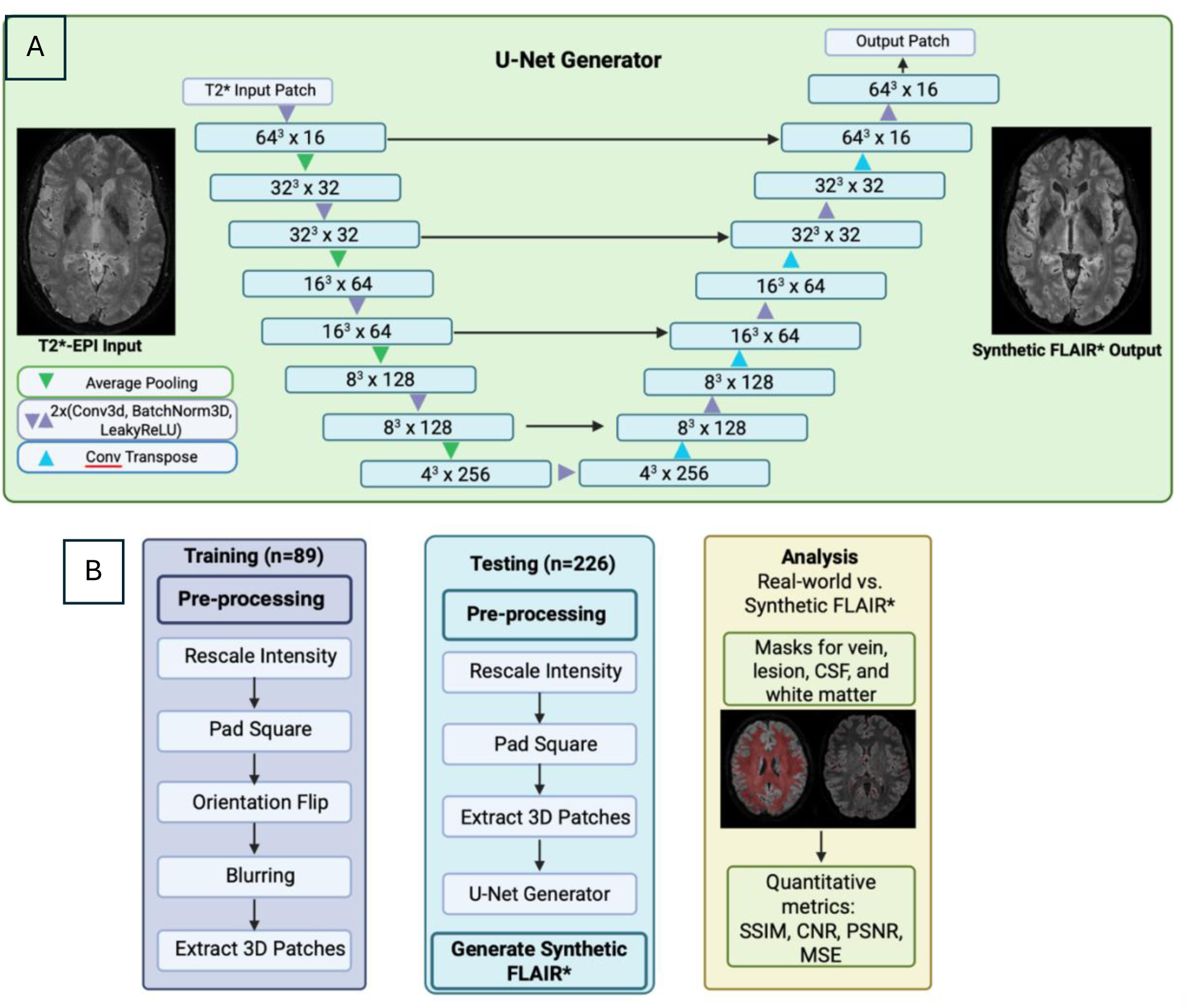
(A) Architecture of U-Net generator used to transform a T2*-EPI input image to a synthetic FLAIR* output image. The encoder consisted of four convolutional stages, each comprising a 3D convolution, batch normalization, and LeakyReLU activation layers, followed by 3D average-pooling layers. Feature dimensionality increased across scales (16, 32, 64, and 128 channels), with a 256-channel bottleneck representation. The decoder mirrored this structure, employing 3D transposed convolutions for upsampling and symmetric skip-connections between encoder and decoder stages. Each decoding stage applied a convolution, batch normalization, and LeakyReLU block. A final 1×1×1 convolution layer projected to a single output channel, and ReLU activation was applied to constrain voxel intensities to non-negative values. (B) Pre- and post-processing steps used during model training and testing. For intensity rescaling, T2* 3D EPI images were rescaled to the range [0,1]. Images were padded to size 320×384×320, flipped (horizontally, vertically, or both), optional blurring was performed, and patches of size 64×64×64 were extracted with a stride of 32 along each axis. SSIM=structural similarity index measures, CNR=contrast-to-noise ratio, PSNR=peak signal-to-noise ratio, MSE=mean squared error.

Hyperparameter optimization was performed using Optuna, with a search space including learning rate [10⁻⁵, 10⁻³], Adam optimizer parameters β₁ [0.4, 0.9] and β₂ [0.9, 0.999], and batch size {4, 8, 16, 32}^21^. Model selection was based on minimizing validation loss over 75 epochs using a composite objective function combining voxel wise mean-squared error (MSE), a perceptual similarity term (1−SSIM), and a directional finite-difference gradient consistency term averaged across each axis. Gradient clipping (norm ≤ 1.0) was applied during training optimization and the best-performing hyperparameter configuration was retrained on the full training set for 75 epochs.

For model testing and evaluation, brain volumes were preprocessed and passed through the frozen generator. Output patches were reassembled using a 3D Hann overlap-add window with per-voxel weight normalization to suppress seam artifacts. Padding from the reconstructed volumes was then removed to native dimensions and the images were saved as NIfTI files. Synthetic FLAIR* images were compared to real-world FLAIR* images which were obtained by standard post-processing of paired T2-FLAIR and T2* acquisitions. All experiments were conducted on an Ubuntu workstation with 32 cores 3.7 GHz CPUs, 256 GB memory, and four NVIDIA Quadro RTX 6000 GPUs. Models were implemented in PyTorch (v2.1.2).

### Quantitative Analysis

Whole-brain image quality was evaluated using structural similarity index measures (SSIM), peak signal-to-noise ratio (PSNR), and mean squared error (MSE) metrics comparing synthetic FLAIR* generated from the DeepFLAIR* model against real-world FLAIR*. Contrast-to-noise ratio (CNR) was computed at lesion-to-vein, lesion-to-white-matter, vein-to-white-matter, and cerebrospinal-fluid-to-white-matter interfaces to quantify small-structure conspicuity relevant to central vein sign evaluation.

White matter, cerebrospinal fluid, and lesion masks were derived from FreeSurfer segmentations on T1 MPRAGE and registered to each subject’s FLAIR* space. Masks were visually inspected for misregistration^22,23^. Vein masks were generated using a Frangi filter^24^.

### Statistical Analysis

All comparisons were conducted as within-subject paired analyses between harmonized real-world FLAIR* and synthetic FLAIR*. Tissue-specific harmonization was performed using ComBat method to correct intensity variations between real-world and synthetic FLAIR* scans prior to pairwise testing^25^. Paired two-sided Wilcoxon signed-rank tests were used to compare each metric between the synthetic and real-world FLAIR* images. Multiple comparisons were controlled using the Benjamini–Hochberg false discovery rate (FDR) at α = 0.05. Tissue-specific intensity harmonization and region-of-interest extraction were implemented in MATLAB, and statistical testing and visualization were performed in Python (NumPy, SciPy, Matplotlib).

## RESULTS

### Visual Evaluation

**Figure 2** illustrates a a representative MS lesion across multiple modalities, including T2-FLAIR, T2*3D-EPI, real-world FLAIR*, and the synthetic FLAIR* image. The magnified panel confirmed that the presence of a CVS positive lesion remains well defined following processing with DeepFLAIR*. **Figure 3** shows axial and coronal views of real-world and synthetic FLAIR* images, where the same CVS-positive MS lesion was identifiable in both images. Cortical boundaries, however, appeared slightly less sharply defined in some regions. Across subjects, lesion geometry, perivenular orientation, and lesion–vein spatial relationship was consistently preserved, indicating that synthetic FLAIR* maintained disease-relevant CVS signatures.

**Figure 2:**
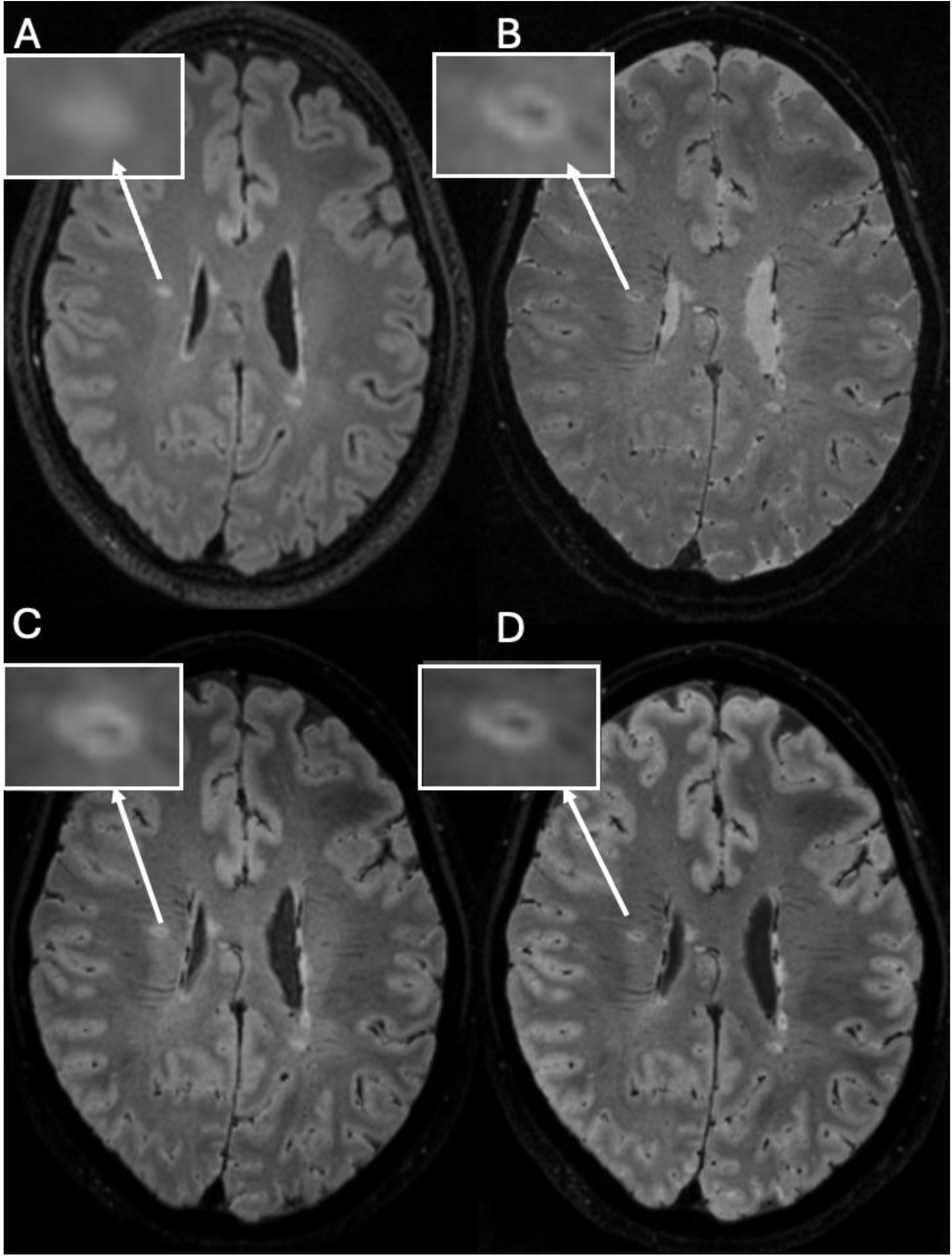
(A) T2-FLAIR, (B) T2* 3D-EPI, (C) Real-world FLAIR*, (D) Synthetic FLAIR*. Magnified panel shows CVS positive MS lesion. CVS=central vein sign, MS=multiple sclerosis.

**Figure 3:**
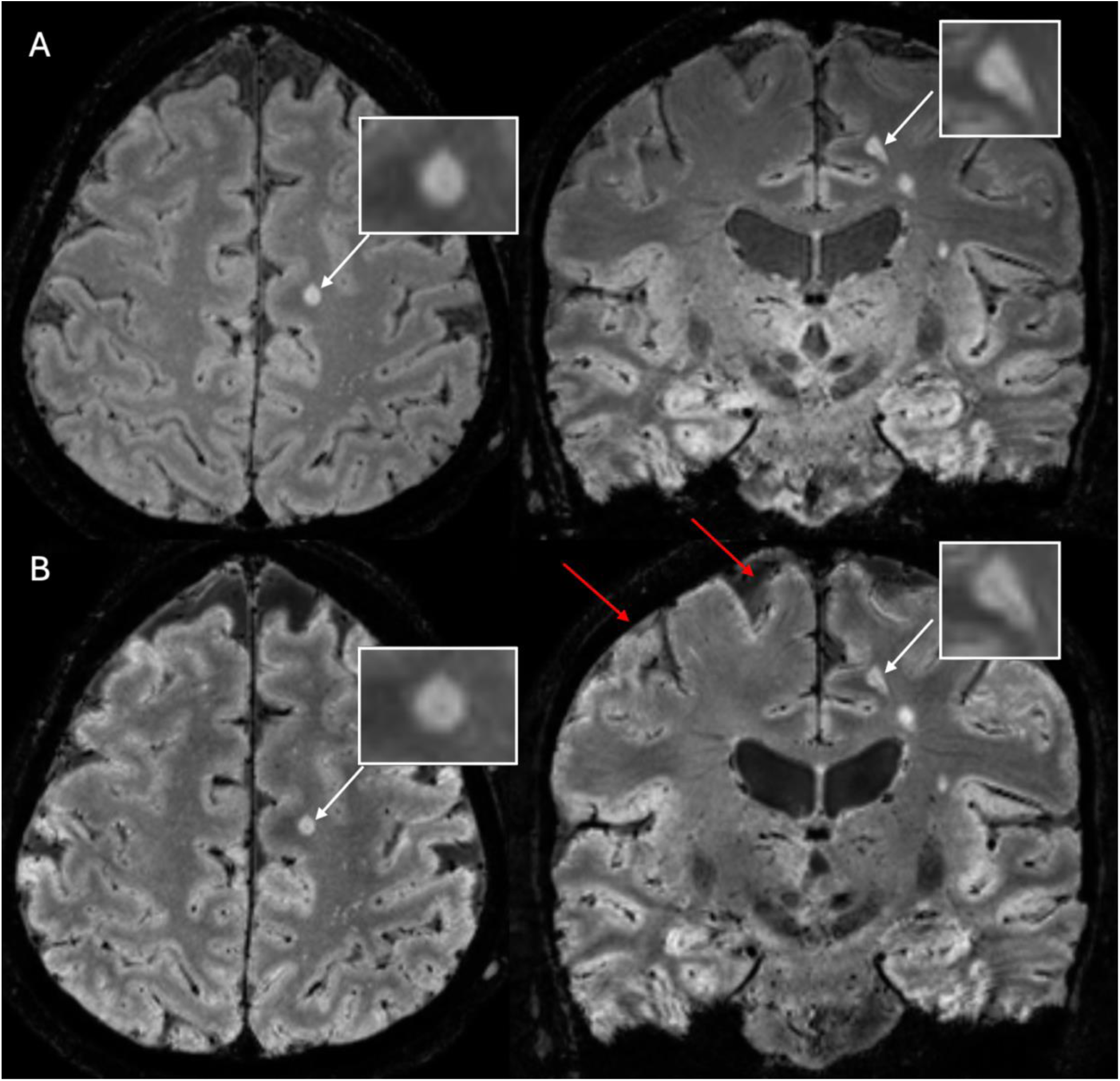
(A) Real-world FLAIR* in axial view (left) and coronal view (right). (B) Synthetic FLAIR* in axial view (left) and coronal view (right). White arrows point to the same CVS positive MS lesion in all four slices. Red arrows point at areas that need improvement in the synthetic FLAIR*. CVS=central vein sign, MS=multiple sclerosis.

### Comparison of Quantitative Metrics

Whole-brain image quality metrics further supported anatomical fidelity between harmonized real-world and synthetic FLAIR*. The synthetic images demonstrated high structural correspondence to the real-world images (mean SSIM = 0.78 ± 0.03), indicating consistent preservation of major anatomical and lesion boundaries. Voxel-wise intensity differences remained low (MSE = 0.0045 ± 0.0015), and PSNR measured 23.6 ± 1.35 dB, supporting consistent global image appearance between the two domains.

Contrast-to-noise ratios (CNR) were evaluated across four clinically relevant tissue interfaces: lesion–vein, lesion–white matter, vein–white matter, and white matter–CSF. As shown in **Figure 4**, synthetic FLAIR* exhibited higher CNR in the lesion–vein, vein–white matter, and white matter–CSF interfaces compared to harmonized real-world FLAIR*, indicating sharper delineation of vessel boundaries and perivascular lesion contrast. These effects were highly consistent across subjects, with all paired comparisons remaining significant following outlier removal and multiple comparison correction (Wilcoxon; *P* < 0.001).

**Figure 4:**
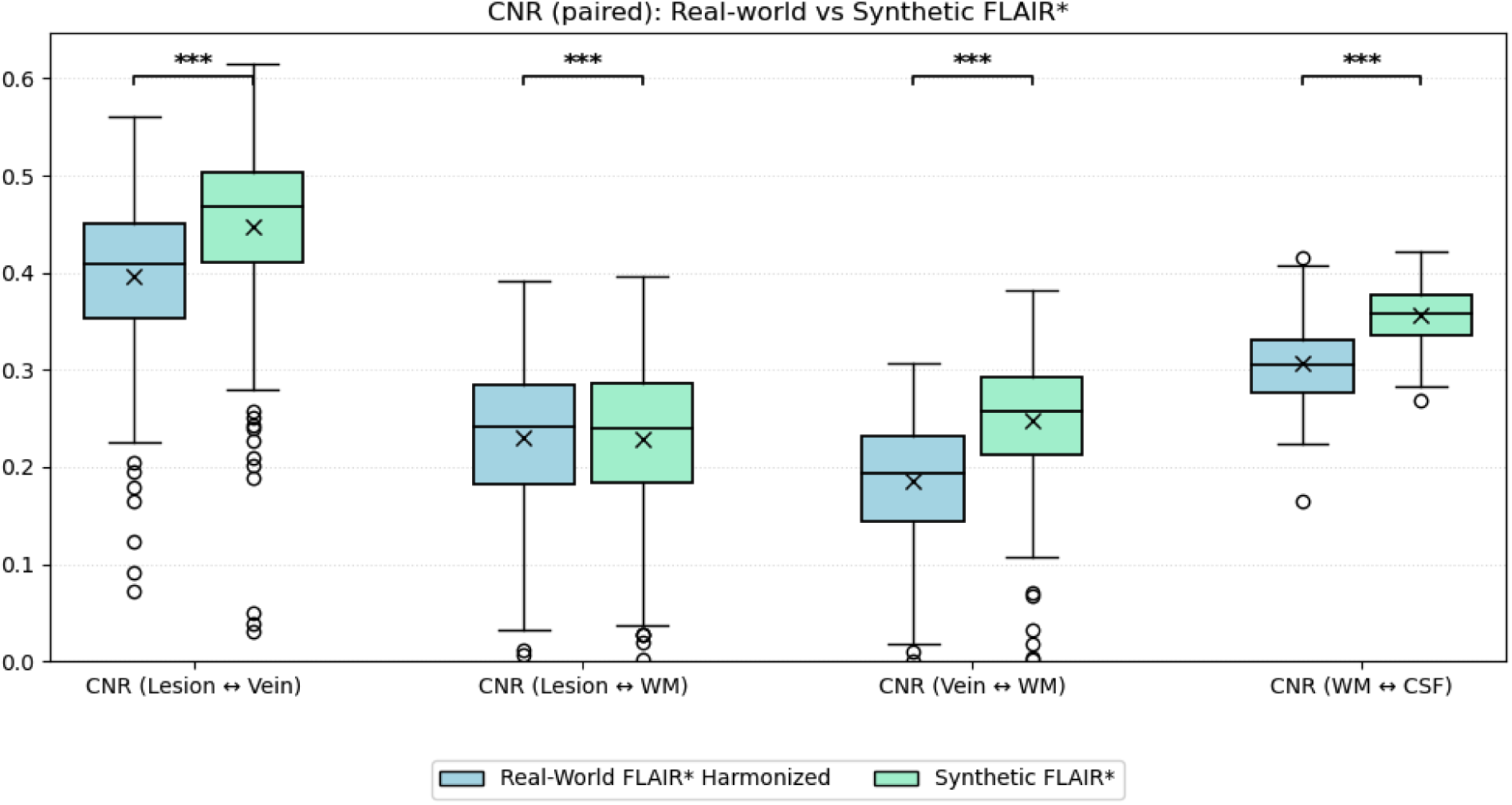
Boxplot depicting the ranges of CNR values for lesion-vein, lesion-white matter, vein-white matter, and cerebrospinal fluid-white matter when comparing real-world and synthetic FLAIR*. Statistically significant differences indicated using asterisks (P*** < 0.001).

## DISCUSSION

This study introduces and validates DeepFLAIR* as a feasible deep learning–based method for synthesizing diagnostically useful FLAIR* contrast directly from T2* 3D EPI MRI, removing the need for dual-sequence acquisition. Across both global similarity metrics and focal lesion-based contrast analyses, synthetic FLAIR* closely matched real-world FLAIR* while preserving perivenular lesion architecture critical for CVS evaluation. In particular, lesion-vein interfaces in synthetic FLAIR* images demonstrated consistently elevated contrast-to-noise ratios, suggesting enhanced visibility of perivenular structure. This pattern reflects a stable, model-learned contrast profile, rather than random or subject-specific variation, that improves visibility of lesion-vascular structure relationships relevant to central vein sign assessment. Together, these findings support the feasibility of enabling CVS-based diagnostic assessment without requiring dual-sequence acquisition or offline image fusion.

In comparison, prior work in synthetic susceptibility imaging, including DeepSWI, has focused primarily on enhancing venous and microbleed contrast from SWI magnitude images^20^. While these studies demonstrated that deep learning can recover susceptibility-driven signal loss and microvascular architecture, they were not designed to reproduce the combined lesion-suppressed and CSF-suppressed contrast profile required for FLAIR*. In contrast, DeepFLAIR* represents the first model to synthesize FLAIR* contrast directly and is optimized to preserve the lesion–vein spatial relationship that defines the central vein sign. Accordingly, our model addresses a distinct diagnostic objective—enabling direct visualization of perivenular lesions as a structural biomarker for multiple sclerosis.

Despite these strengths, several limitations should be acknowledged. First, cortical boundaries appeared slightly less sharp, suggesting that additional optimization may be needed to better preserve fine structural detail in these regions. Additionally, CSF suppression near the skull in synthetic FLAIR* occasionally appeared less uniform compared to real-world FLAIR*, suggesting an opportunity to refine intensity calibration near tissue-CSF interfaces. These limitations may benefit from incorporating a discriminator component into the network architecture to further refine fine-scale structural detail and improve intensity consistency across tissue boundaries. Furthermore, model training was performed exclusively on 3T Siemens and Philips systems using standardized acquisition protocols; generalizability to 1.5T systems or higher-field (7T) acquisitions remains to be validated. Finally, although synthetic FLAIR* images preserved CVS morphology in qualitative and CNR-based analyses, expert diagnostic grading was not performed in this study.

Addressing these limitations, future work will focus on refining network architecture and training objectives to enhance cortical sharpness and achieve more uniform CSF suppression, potentially through the inclusion of a discriminator-based adversarial network to better capture fine-scale contrast variations. Blinded neuroradiologist scoring of CVS visibility in paired real-world and synthetic FLAIR* images will further assess diagnostic interchangeability and evaluate potential false positive or false negative lesion classifications. Incorporating a wide range of MRI vendors, acquisition parameters for T2* 3D EPI contrasts, and patient populations will further support model robustness and clinical deployment. In addition, synthetic FLAIR* provides a promising foundation for automated CVS detection pipelines, which could enable fully integrated biomarker-based MS evaluation using a single acquisition sequence, reducing scan time and post-processing demands in routine practice. Collectively, these directions aim to establish DeepFLAIR* as a scalable clinical tool that maintains diagnostic integrity across scanners, populations, and institutions.

In conclusion, DeepFLAIR* offers a clinically practical strategy for generating diagnostically meaningful FLAIR* contrast from T2* 3D EPI data. By eliminating the need for dual-sequence acquisition and offline fusion, this approach has the potential to streamline imaging workflows, increase CVS accessibility, and support more accurate, rapid MS diagnosis in real-world settings.

## Data Availability Statement

The CAVS-MS data used in this study may be available upon reasonable request from co-investigators P.S., N.S., and D.O. through a formal data sharing agreement.

This model is owned by and proprietary to Cedars-Sinai Medical Center. © 2025 Cedars-Sinai Medical Center. All rights reserved. For any use requests, please reach out to.

## Clinical Trial and Ethics Statement

IRB of Cleveland Clinic gave ethical approval of this work.

https://ClinicalTrials.gov ID: NCT04495556.

## REFERENCES

1. Wallin MT, Culpepper WJ, Campbell JD, Nelson LM, Langer-Gould A, Marrie RA, Cutter GR, Kaye WE, Wagner L, Tremlett H, Buka SL, Dilokthornsakul P, Topol B, Chen LH, LaRocca NG, US Multiple Sclerosis Prevalence Workgroup. The prevalence of MS in the United States: A population-based estimate using health claims data. Neurology. 2019 Mar 5;92(10):e1029–e1040. PMCID: PMC6442006

2. Dilokthornsakul P, Valuck RJ, Nair KV, Corboy JR, Allen RR, Campbell JD. Multiple sclerosis prevalence in the United States commercially insured population. Neurology. 2016 Mar 15;86(11):1014–1021. PMCID: PMC4799713

3. Reich DS, Lucchinetti CF, Calabresi PA. Multiple Sclerosis. N Engl J Med. 2018 Jan 11;378(2):169–180. PMCID: PMC6942519

4. Thompson AJ, Banwell BL, Barkhof F, Carroll WM, Coetzee T, Comi G, Correale J, Fazekas F, Filippi M, Freedman MS, Fujihara K, Galetta SL, Hartung HP, Kappos L, Lublin FD, Marrie RA, Miller AE, Miller DH, Montalban X, Mowry EM, Cohen JA. Diagnosis of multiple sclerosis: 2017 revisions of the McDonald criteria. Lancet Neurol. 2018 Feb;17(2):162–173. PMID: 29275977

5. Filippi M, Rocca MA, De Stefano N, Enzinger C, Fisher E, Horsfield MA, Inglese M, Pelletier D, Comi G. Magnetic resonance techniques in multiple sclerosis: the present and the future. Arch Neurol. 2011 Dec;68(12):1514–1520. PMID: 22159052

6. Kaisey M, Solomon AJ, Luu M, Giesser BS, Sicotte NL. Incidence of multiple sclerosis misdiagnosis in referrals to two academic centers. Mult Scler Relat Disord. 2019 May;30:51–56. PMID: 30738280

7. Sati P, Oh J, Constable RT, Evangelou N, Guttmann CRG, Henry RG, Klawiter EC, Mainero C, Massacesi L, McFarland H, Nelson F, Ontaneda D, Rauscher A, Rooney WD, Samaraweera APR, Shinohara RT, Sobel RA, Solomon AJ, Treaba CA, Wuerfel J, NAIMS Cooperative. The central vein sign and its clinical evaluation for the diagnosis of multiple sclerosis: a consensus statement from the North American Imaging in Multiple Sclerosis Cooperative. Nat Rev Neurol. 2016 Dec;12(12):714–722. PMID: 27834394

8. Daboul L, O’Donnell CM, Cao Q, Amin M, Rodrigues P, Derbyshire J, Azevedo C, Bar-Or A, Caverzasi E, Calabresi P, Cree BAC, Freeman L, Henry RG, Longbrake EE, Nakamura K, Oh J, Papinutto N, Pelletier D, Samudralwar RD, Suthiphosuwan S, Sati P. Effect of GBCA Use on Detection and Diagnostic Performance of the Central Vein Sign: Evaluation Using a 3-T FLAIR* Sequence in Patients With Suspected Multiple Sclerosis. AJR Am J Roentgenol. 2023 Jan;220(1):115–125. PMCID: PMC10016223

9. Montalban X, Lebrun-Frénay C, Oh J, Arrambide G, Moccia M, Pia Amato M, Amezcua L, Banwell B, Bar-Or A, Barkhof F, Butzkueven H, Ciccarelli O, Chataway J, Cohen JA, Comi G, Correale J, Deisenhammer F, Filippi M, Fiol J, Freedman MS, Fujihara K, Granziera C, Green AJ, Hartung HP, Hellwig K, Kappos L, Kimbrough D, Killestein J, Lublin F, Marignier R, Ann Marrie R, Miller A, Otero-Romero S, Ontaneda D, Ramanathan S, Reich D, Rocca MA, Rovira À, Saidha S, Salter A, Sastre-Garriga J, Saylor D, Solomon AJ, Sormani MP, Stankoff B, Tintore M, Tremlett H, Van der Walt A, Viswanathan S, Wiendl H, Wildemann B, Yamout B, Zaratin P, Calabresi PA, Coetzee T, Thompson AJ. Diagnosis of multiple sclerosis: 2024 revisions of the McDonald criteria. Lancet Neurol. 2025 Oct;24(10):850–865. PMID: 40975101.

10. Sati P, George IC, Shea CD, Gaitán MI, Reich DS. FLAIR*: a combined MR contrast technique for visualizing white matter lesions and parenchymal veins. Radiology. 2012 Dec;265(3):926–932. PMCID: PMC3504317

11. Sati P, Patil S, Inati S, Wang WT, Derbyshire JA, Krueger G, Reich DS, Butman JA. Rapid MR susceptibility imaging of the brain using segmented 3D echo-planar imaging and its clinical applications. MAGNETOM Flash. 2017;(68):26.

12. Solomon AJ, Schindler MK, Howard DB, Watts R, Sati P, Nickerson JP, Reich DS. “Central vessel sign” on 3T FLAIR* MRI for the differentiation of multiple sclerosis from migraine. Ann Clin Transl Neurol. 2016 Feb;3(2):82–87. PMCID: PMC4748312

13. Kilsdonk ID, Wattjes MP, Lopez-Soriano A, Kuijer JPA, de Jong MC, de Graaf WL, Conijn MMA, Polman CH, Luijten PR, Geurts JJG, Geerlings MI, Barkhof F. Improved differentiation between MS and vascular brain lesions using FLAIR* at 7 Tesla. Eur Radiol. 2014 Apr;24(4):841–849. PMID: 24317461

14. Daboul L, O’Donnell CM, Amin M, Rodrigues P, Derbyshire J, Azevedo C, Bar-Or A, Caverzasi E, Calabresi PA, Cree BA, Freeman L, Henry RG, Longbrake EE, Oh J, Papinutto N, Pelletier D, Prchkovska V, Raza P, Ramos M, Samudralwar RD, Ontaneda D. A multicenter pilot study evaluating simplified central vein assessment for the diagnosis of multiple sclerosis. Mult Scler. 2024 Jan;30(1):25–34. PMCID: PMC11037932

15. Toljan K, Daboul L, Raza P, Martin ML, Cao Q, O’Donnell CM, Rodrigues P, Derbyshire J, Azevedo CJ, Bar-Or A, Caverzasi E, Calabresi PA, Cree BA, Freeman L, Henry RG, Longbrake EE, Oh J, Papinutto N, Pelletier D, Samudralwar RD, Schindler MK, Sotirchos ES, Sicotte NL, Solomon AJ, Shinohara RT, Reich DS, Sati P, Ontaneda D. Diagnostic performance of central vein sign versus oligoclonal bands for multiple sclerosis. Mult Scler. 2024 Sep;30(10):1268–1277. doi: 10.1177/13524585241271988. Epub 2024 Sep 5. PMCID: PMC11421977.

16. Manning AR, Letchuman V, Martin ML, Gombos E, Robert-Fitzgerald T, Cao Q, Raza P, O’Donnell CM, Renner B, Daboul L, Rodrigues P, Ramos M, Derbyshire J, Azevedo C, Bar-Or A, Caverzasi E, Calabresi PA, Cree BAC, Freeman L, Henry RG, Longbrake EE, Oh J, Papinutto N, Pelletier D, Samudralwar RD, Suthiphosuwan S, Schindler MK, Bilello M, Song JW, Sotirchos ES, Sicotte NL, Al-Louzi O, Solomon AJ, Reich DS, Ontaneda D, Sati P, Shinohara RT; NAIMS Cooperative. Multicenter Automated Central Vein Sign Detection Performs as Well as Manual Assessment for the Diagnosis of Multiple Sclerosis. AJNR Am J Neuroradiol. 2025 Mar 4;46(3):620–626. doi: 10.3174/ajnr.A8510. PMCID: PMC11979820.

17. Amin M, Nakamura K, Daboul L, O’Donnell C, Cao Q, Rodrigues P, Derbyshire J, Azevedo C, Bar-Or A, Caverzasi E, Calabresi PA, Cree BAC, Freeman L, Henry R, Longbrake EE, Oh J, Papinutto N, Pelletier D, Prčkovska V, Raza PC, Ramos M, Samudralwar R, Schindler M, Sotirchos ES, Sicotte N, Solomon AJ, Shinohara R, Reich DS, Sati P, Ontaneda D. Incorporation of the central vein sign into the McDonald criteria. Mult Scler Relat Disord. 2025 Jan;93:106182. doi: 10.1016/j.msard.2024.106182. Epub 2024 Nov 25. PMCID: PMC11779579.

18. Campion T, Smith RJP, Altmann DR, Brito GC, Turner BP, Evanson J, George IC, Sati P, Reich DS, Miquel ME, Schmierer K. FLAIR* to visualize veins in white matter lesions: A new tool for the diagnosis of multiple sclerosis? Eur Radiol. 2017 Oct;27(10):4257–4263. doi: 10.1007/s00330-017-4822-z. Epub 2017 Apr 13. PMCID: PMC5579202.

19. Ontaneda D, Sati P, Raza P, Kilbane M, Gombos E, Alvarez E, Azevedo C, Calabresi P, Cohen JA, Freeman L, Henry RG, Longbrake EE, Mitra N, Illenberger N, Schindler M, Moreno-Dominguez D, Ramos M, Mowry E, Oh J, Rodrigues P, North American Imaging in MS Cooperative. Central vein sign: A diagnostic biomarker in multiple sclerosis (CAVS-MS) study protocol for a prospective multicenter trial. Neuroimage Clin. 2021 Sep 23;32:102834. PMCID: PMC8482479

20. Genc O, Morrison MA, Villanueva-Meyer JE, Burns B, Hess CP, Banerjee S, Lupo JM. DeepSWI: Using Deep Learning to Enhance Susceptibility Contrast on T2*-Weighted MRI. J Magn Reson Imaging. 2023 Oct;58(4):1200–1210. PMCID: PMC10443940

21. Akiba T, Sano S, Yanase T, Ohta T, Koyama M. Optuna: A Next-generation Hyperparameter Optimization Framework. arXiv [Preprint] 2019 Jul 25:1907.10902. doi:10.48550/arXiv.1907.10902.

22. Fischl B. FreeSurfer. Neuroimage. 2012 Aug 15;62(2):774–81. PMCID: PMC3685476.

23. Al-Louzi O, Letchuman V, Manukyan S, Beck ES, Roy S, Ohayon J, Pham DL, Cortese I, Sati P, Reich DS. Central Vein Sign Profile of Newly Developing Lesions in Multiple Sclerosis: A 3-Year Longitudinal Study. Neurol Neuroimmunol Neuroinflamm. 2022 Jan 13;9(2):e1120. PMCID: PMC8759076.

24. Frangi AF, Niessen WJ, Vincken KL, Viergever MA. Multiscale vessel enhancement filtering. Medical image computing and computer-assisted intervention—MICCAI’98: first international conference cambridge, MA, USA, october 11–13, 1998 proceedings 1; 1998: Springer.

25. Fortin JP, Cullen N, Sheline YI, Taylor WD, Aselcioglu I, Cook PA, Adams P, Cooper C, Fava M, McGrath PJ, McInnis M, Phillips ML, Trivedi MH, Weissman MM, Shinohara RT. Harmonization of cortical thickness measurements across scanners and sites. Neuroimage. 2018 Feb 15;167:104–120. PMCID: PMC5845848.

